## Supplementary Material for "Seroprevalence of SARS-CoV-2 antibodies and retrospective mortality in a refugee camp, Dagahaley, Kenya"

- 1.3 Seroprevalence standardized by age and sex and adjusted for sensitivity and specificity of the test by recruitment status 5

### Tables

### Figures

### 1 Seroprevalence

#### 1.1 Crude TDR Results by sex

Males had slightly higher seroprevalence than female, which was not statistically significant.

Table 1. Results of Rapid Diagnostic Test BIOSYNEX COVID-19 BSS by sex and overall, Dagahaley refugee camp, Garissa County, Kenya, May 2021

| Age group | Negative RDT | IgG + | IgM+ | IgG+ & IgM+ | Positive (%) | 95% CI |
|---|---|---|---|---|---|---|
| Female | 621 | 24 | 15 | 12 | 7.6 | [5.6-9.6] |
| Male | 488 | 26 | 13 | 7 | 8.6 | [6.2-11] |
| Total | 1109 | 50 | 28 | 19 | | |

#### 1.2 Crude TDR Results by age group

The lower proportion of positive to the test was found in children under 5 years, the higher among people aged 20 to 34 years and people 50 years or above.  As shown in figure 3, the proportion increase until the age of 25 to stabilize above.

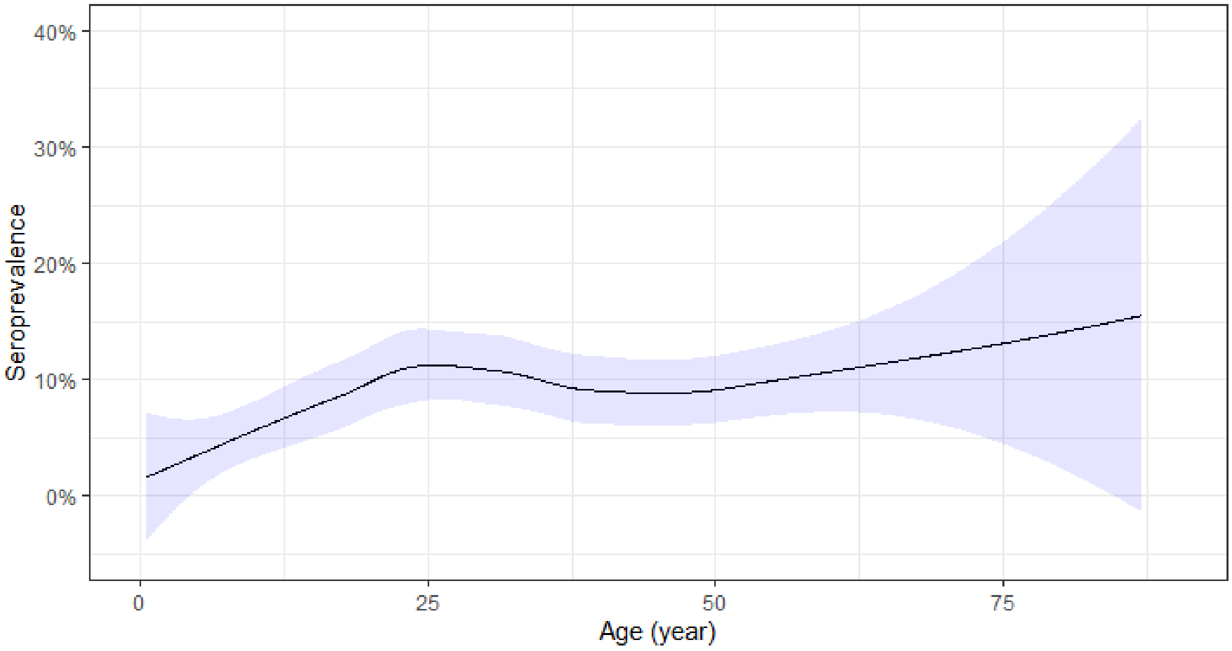

Figure 1 Proportion of seropositive by age (positive either IgG or IgM by Rapid Diagnostic Test BIOSYNEX COVID-19 BSS ), Dagahaley refugee camp, Garissa County, Kenya, May 2021

#### 1.3 Seroprevalence standardized by age and sex and adjusted for sensitivity and specificity of the test by recruitment status

Table 2. Seroprevalence standardized by age and sex and adjusted for sensitivity and specificity of the test (Rapid Diagnostic Test ) by mode of recruitment, Dagahaley refugee camp, Garissa County, Kenya, May 2021

| | Crude | Standardized by age | Adjusted for test performance | 95% CI |
|---|---|---|---|---|
| CHW, TBA and their family members | 9.2 % | 8.8 % | 7.5% | [2.4-11.1] |
| patient and caretaker | 6.9 % | 3.9 % | 2.8% | [0.3-5.2] |
| Overall | 8 % | 7.2 % | **5.8%** | **[1.6-8.4]** |

#### 1.4 Risk factor associated with seroconversion

We explored the risk factors associated with seropositivity through a multivariate logistic regression. Being aged 20 years or more was significantly associated with an increase risk, sex was not significantly associated. Being a family members of a community health worker or a traditional birth Attendant was associated with an increased risk (RR=1.9), being a CHW or a TBA was also associated to an increased risk though not statistically significant which can be due to the lower sample size of this group.

Table 3. Multivariate analysis of risk factors associated with seropositivity , Dagahaley refugee camp, Garissa County, Kenya, May 2021

| | N | Relative Risk | 95% CI | P value |
|---|---|---|---|---|
| 0-19 Y | 510 | reference | | |
| 20-34 Y | 261 | **2.81** | [1.54-5.11] | <0.001 |
| 35-49 Y | 231 | **2.1** | [1.01-4.3] | 0.045 |
| >=50 Y | 204 | **3.26** | [1.65-6.41] | <0.001 |
| Sex (Female) | 672 | reference | | |
| Sex (male) | 534 | **0.82** | [0.55-1.24] | 0.3503 |
| Patient or caretaker | 619 | reference | | |
| Community Health Worker or Traditional Birth Attendant | 145 | **1.57** | [0.88-2.7] | 0.117 |
| Family member of CHW or TBA | 442 | **1.88** | [1.08-3.23] | 0.023 |

The recruitment of the whole Households of CHW and TBA, allows us to explore the risk factor associated with having another person seropositive in the households. The analysis in this subgroup shows that this is significantly associated with an increase more than two folds higher. In contrast the size of the household doesn't seems to increase the risk.

Table 4. Sub-analysis among Household of community health worker and TBA of risk factor associated with seroconversion, Dagahaley refugee camp, Garissa County, Kenya, May 2021

| | N | Relative Risk | 95% CI | P value |
|---|---|---|---|---|
| 0-19 Years | 354 | reference | | |
| >=20 Years | 233 | **2.87** | [1.54-5.46] | <0.001 |
| Male | 302 | reference | | |
| Female | 302 | **0.78** | [0.42-1.42] | 0.402 |
| Household size | 587 | **0.98** | [0.82-1.17] | 0.819 |
| not exposed to another seropositive in HH | 551 | reference | | |
| exposed to another seropositive in HH | 36 | **2.69** | [1.37-5.18] | 0.0022 |

#### 1.5 Morbidities and health seeking behaviour

The dates of onset of symptoms are consistent with the peak of reported cases in Dagahaley in September 2020.

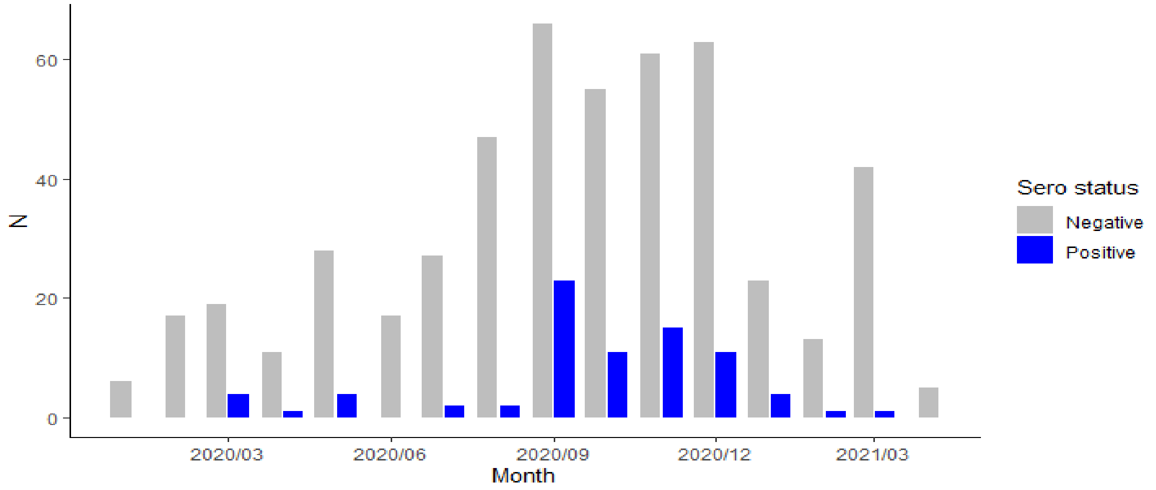

Figure 2. Date of onset of symptoms by Results COVID-19 seropositivity status, Dagahaley refugee camp, Garissa County, Kenya, May 2021.

Table 5 shows the symptoms reported according to seropositivity status. Almost all of them (but nausea, vomiting, abdominal pain, diarrhoea and altered mental status) were significantly more frequent in seropositive participants.

Table 5. Symptoms associated with COVID-19 by Results of Rapid Diagnostic Test BIOSYNEX COVID-19 BSS, Dagahaley refugee camp, Garissa County, Kenya, May 2021

| | Negative | Positive | P value* | IgM + | IgG+ & IgM+ | IgG + |
|---|---|---|---|---|---|---|
| Fever | 21% (236) | 63% (61) | 0 | 50% (14) | 79% (15) | 64% (32) |
| Chills | 5% (55) | 27% (26) | 0 | 21% (6) | 21% (4) | 32% (16) |
| Fatigue | 9% (96) | 40% (39) | 0 | 29% (8) | 37% (7) | 48% (24) |
| Myalgia | 11% (125) | 40% (39) | 0 | 39% (11) | 42% (8) | 40% (20) |
| Sore throat | 7% (73) | 34% (33) | 0 | 32% (9) | 26% (5) | 38% (19) |
| Cough | 27% (299) | 70% (68) | 0 | 68% (19) | 84% (16) | 66% (33) |
| Rhinorrhea | 17% (183) | 49% (47) | 0 | 46% (13) | 53% (10) | 49% (24) |
| Shortness of breath | 3% (31) | 11% (11) | 0 | 11% (3) | 11% (2) | 12% (6) |
| Wheezing | 1% (11) | 9% (9) | 0 | 11% (3) | 11% (2) | 8% (4) |
| Chest pain | 9% (103) | 33% (32) | 0 | 25% (7) | 37% (7) | 36% (18) |
| Other respiratory symptoms | 1% (14) | 15% (15) | 0 | 4% (1) | 16% (3) | 22% (11) |
| Headache | 29% (315) | 64% (62) | 0 | 71% (20) | 63% (12) | 60% (30) |
| Nausea vomiting | 4% (43) | 8% (8) | 0.059 | 4% (1) | 5% (1) | 12% (6) |
| Abdominal pain | 6% (68) | 12% (12) | 0.03 | 11% (3) | 21% (4) | 10% (5) |
| Diarrhea | 3% (33) | 6% (6) | 0.123 | 4% (1) | 11% (2) | 6% (3) |
| Loss of taste | 6% (66) | 33% (32) | 0 | 29% (8) | 26% (5) | 38% (19) |
| Loss of smell | 5% (53) | 26% (25) | 0 | 25% (7) | 26% (5) | 26% (13) |
| Altered mental status | 1% (8) | 2% (2) | 0.187 | 0% (0) | 0% (0) | 4% (2) |

***Pvalue is comparing participants negatives vs positive***

#### 1.6 Comorbidities

Comorbidities were similar in participants who tested positive and negative. The most common comorbidity was hypertension.

Table 6. Comorbidities and medical conditions among participants aged 18 years or more by results of Rapid Diagnostic Test BIOSYNEX COVID-19 BSS, Dagahaley refugee camp, Garissa County, Kenya, May 2021

| | Positive RDT | | Negative RDT | |
|---|---|---|---|---|
| | Yes | Unknown | Yes | Unknown |
| Hypertension | 11% (8/75) | 0% (n=0) | 9% (61/668) | 1% (n=8) |
| Asthma | 3% (2/75) | 0% (n=0) | 2% (13/674) | 0% (n=2) |
| Autoimmune Disease | 0% (0/75) | 0% (n=0) | 0% (0/673) | 0% (n=3) |
| Cancer | 0% (0/75) | 0% (n=0) | 0% (1/669) | 1% (n=7) |
| Chronic Lung Disease | 0% (0/74) | 1% (n=1) | 0% (3/672) | 1% (n=4) |
| Congestive Heart Failure | 0% (0/73) | 3% (n=2) | 1% (5/675) | 0% (n=1) |
| Coronary Heart Disease | 0% (0/75) | 0% (n=0) | 1% (8/675) | 0% (n=1) |
| Diabetes | 8% (6/75) | 0% (n=0) | 3% (22/656) | 3% (n=20) |
| Hepatitis B | 1% (1/75) | 0% (n=0) | 1% (9/668) | 1% (n=8) |
| Hepatitis C | 0% (0/75) | 0% (n=0) | 1% (4/671) | 1% (n=5) |
| Obesity | 1% (1/74) | 1% (n=1) | 0% (2/673) | 0% (n=3) |
| Past Transplant | 0% (0/75) | 0% (n=0) | 0% (1/672) | 1% (n=4) |
| HIV | 0% (0/72) | 4% (n=3) | 0% (2/646) | 4% (n=30) |
| Kidney Disease | 11% (8/75) | 0% (n=0) | 7% (45/670) | 1% (n=6) |
| Cirhosis | 0% (0/73) | 3% (n=2) | 1% (7/657) | 3% (n=19) |
| Smoker (current or former) | 4% (3/75) | 0% (n=0) | 3% (20/676) | 0% (n=0) |

### 2 Mortality

#### 2.1 Death by age group and by months

According to the literature COVID-19 Infectious Fatality Rate is much higher in people over 50 years. To explore if an excess mortality is higher in this age group as a direct consequence of the pandemic we estimated the proportion of death over 50 years over time (figure 3 ) but identified no significant trend.

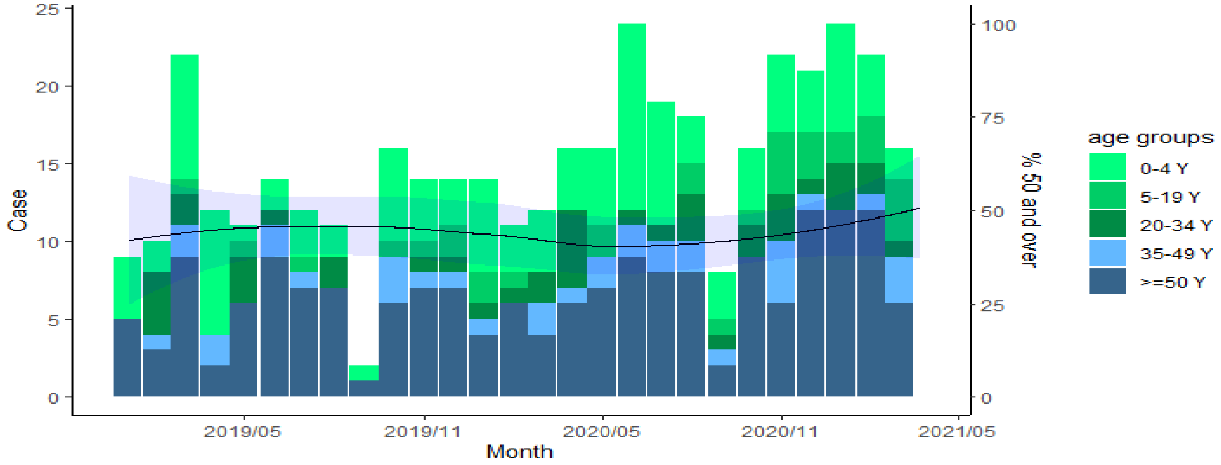

Figure 3. Number of deaths by age group and by month and Local polynomial Regression of the proportion of death over 50, Dagahaley refugee camp, Garissa County, Kenya, May 2021

#### 2.2 Estimation of mortality rate before pandemic and during pandemic using the same months of the years

Table 7. Mortality rates by age groups and by period, Dagahaley refugee camp, Garissa County, Kenya

| Age group | Death Pre-Pandemic (May 2019, March 2020) | Mortality rate (per 10 000 pers.day) | 95% CI | Death Pandemic (May 2020, March 2021) | Mortality rate (per 10 000 pers.day) | 95% CI | rate ratio | p value |
|---|---|---|---|---|---|---|---|---|
| 0-4 Y | 33 | 0.07 | [0.05-0.1] | 58 | 0.12 | [0.09-0.15] | 1.62 | 0.026 |
| 5-19 Y | 9 | 0.01 | [0-0.02] | 22 | 0.02 | [0.01-0.03] | 2.25 | 0.036 |
| 20-34 Y | 12 | 0.02 | [0.01-0.04] | 18 | 0.03 | [0.02-0.05] | 1.38 | 0.387 |
| 35-49 Y | 11 | 0.04 | [0.02-0.07] | 18 | 0.06 | [0.04-0.09] | 1.5 | 0.283 |
| >=50 Y | 63 | 0.37 | [0.29-0.47] | 91 | 0.49 | [0.4-0.6] | 1.33 | 0.083 |
| overall | 200 | **0.05** | [0.05-0.06] | 207 | **0.07** | [0.06-0.08] | **1.42** | <0.001 |

#### 2.3 Reported cause of death

The main reported cause of death from a known cause was respiratory disease. COVID-19 was reported for 2 deaths. The only cause of death that has a significant difference between the two period is Accident/trauma/violence.

During the pre-pandemic period, 38 % (75/197) of the deaths were rapid or unexpected which was like the pandemic period at 35% (73/206).
The interviewers had the choice of selecting a cause of death from the respondent's answers in a pre-established list or selecting another cause of death and describing it. After rereading these descriptions, we were able to create new categories and reclassify a significant proportion of these other causes of death, although 42 deaths could not be classified. Nevertheless, among these 42 deaths (Tooth ache kidney pain, Suicide, old age etc…) we did not detect any informative differences between the two periods.

Table 8. Reported cause of death by age group and by period, Dagahaley refugee camp, Garissa County, Kenya

| Cause of death | Under 50 Years | | 50 Years and more | | All ages | | |
|---|---|---|---|---|---|---|---|
| | Pre-Pandemic | Pandemic | Pre-Pandemic | Pandemic | Pre-Pandemic | Pandemic | P value |
| Respiratory Disease | 10 % (11) | 19 % (22) | 12 % (11) | 10 % (9) | 11 % (22) | 15 % (31) | 0.31 |
| Isolated Fever Malaria Anemia | 14 % (15) | 16 % (18) | 9 % (8) | 12 % (11) | 12 % (23) | 14 % (29) | 0.55 |
| Hypertension | 3 % (3) | 5 % (6) | 25 % (22) | 22 % (20) | 12 % (25) | 13 % (26) | 1 |
| Neonatal Death | 20 % (22) | 16 % (18) | 0 % (0) | 0 % (0) | 11 % (22) | 9 % (18) | 0.51 |
| Cancer | 8 % (9) | 3 % (4) | 7 % (6) | 14 % (13) | 8 % (15) | 8 % (17) | 0.85 |
| Diarrhea | 6 % (7) | 6 % (7) | 6 % (5) | 3 % (3) | 6 % (12) | 5 % (10) | 0.67 |
| Accident Trauma Violence | 1 % (1) | 5 % (6) | 0 % (0) | 2 % (2) | 0 % (1) | 4 % (8) | 0.04 |
| Maternal Death | 7 % (8) | 6 % (7) | 1 % (1) | 0 % (0) | 4 % (9) | 3 % (7) | 0.62 |
| Diabetes | 2 % (2) | 1 % (1) | 4 % (4) | 4 % (4) | 3 % (6) | 2 % (5) | 0.76 |
| Heart condition | 0 % (0) | 3 % (3) | 3 % (3) | 0 % (0) | 2 % (3) | 1 % (3) | 1 |
| COVID 19 | 0 % (0) | 1 % (1) | 0 % (0) | 1 % (1) | 0 % (0) | 1 % (2) | 0.5 |
| Other | 12 % (13) | 4 % (5) | 13 % (12) | 13 % (12) | 12 % (25) | 8 % (17) | 0.25 |
| Unknown | 18 % (20) | 16 % (18) | 19 % (17) | 18 % (16) | 18 % (37) | 16 % (34) | 0.70 |

#### 2.4 Reported Comorbidities

The most frequent comorbidity was hypertension. The distribution doesn't suggest any significant difference between pre-pandemic and pandemic period.

Table 9. Reported comorbidities of deceased by period, Dagahaley refugee camp, Garissa County, Kenya

| | Pre-Pandemic | | Pandemic | |
|---|---|---|---|---|
| | Yes | Unknown | Yes | Unknown |
| Hypertension | 23% (46/198) | 1% (n=2) | 18% (38/206) | 0% (n=1) |
| Asthma | 2% (4/197) | 2% (n=3) | 3% (6/206) | 0% (n=1) |
| Autoimmune Disease | 1% (1/197) | 2% (n=3) | 0% (0/206) | 0% (n=1) |
| Cancer | 9% (17/197) | 2% (n=3) | 9% (18/206) | 0% (n=1) |
| Chronic Lung Disease | 4% (8/198) | 1% (n=2) | 2% (4/206) | 0% (n=1) |
| Congestive Heart Failure | 5% (9/196) | 2% (n=4) | 4% (8/205) | 1% (n=2) |
| Coronary Heart Disease | 4% (8/196) | 2% (n=4) | 5% (11/206) | 0% (n=1) |
| Diabetes | 7% (14/198) | 1% (n=2) | 4% (9/206) | 0% (n=1) |
| Hepatitis B | 3% (6/197) | 2% (n=3) | 1% (2/205) | 1% (n=2) |
| Hepatitis C | 1% (1/197) | 2% (n=3) | 0% (1/206) | 0% (n=1) |
| Obesity | 0% (0/197) | 2% (n=3) | 0% (1/205) | 1% (n=2) |
| Past Transplant | 2% (3/197) | 2% (n=3) | 1% (2/206) | 0% (n=1) |
| Smoker | 2% (2/123) | 38% (n=77) | 5% (5/110) | 46% (n=95) |

#### 2.5 Reported Symptom before death by period

The most frequent reported symptoms before death was fever. The distribution doesn't suggest any significant difference between pre pandemic and pandemic period.

Table 10. Reported symptoms before death by period, Dagahaley refugee camp, Garissa County, Kenya

| | Pre-Pandemic | | Pandemic | |
|---|---|---|---|---|
| | Yes | Unknown | Yes | Unknown |
| Fever | 46% (91/199) | 0% (n=1) | 42% (87/206) | 1% (n=3) |
| Shortness of Breath | 20% (40/198) | 1% (n=2) | 22% (45/205) | 1% (n=2) |
| Loss Appetite Vomiting Nausea | 19% (38/199) | 0% (n=1) | 18% (37/206) | 1% (n=2) |
| Cough | 18% (36/198) | 1% (n=2) | 22% (46/206) | 1% (n=2) |
| Weakness Fatigue | 15% (29/197) | 2% (n=3) | 17% (34/205) | 0% (n=1) |
| Headaches | 13% (26/198) | 1% (n=2) | 17% (34/206) | 1% (n=2) |
| Muscle Pain | 11% (21/198) | 1% (n=2) | 12% (24/206) | 1% (n=2) |
| Diarrhea | 8% (16/197) | 2% (n=3) | 5% (11/206) | 0% (n=1) |
| Change in Mental State | 3% (6/197) | 2% (n=3) | 9% (18/206) | 1% (n=2) |
| Sore Throat | 4% (7/198) | 1% (n=2) | 4% (9/206) | 2% (n=4) |
| Loss Taste Odour | 2% (4/197) | 2% (n=3) | 1% (2/205) | 1% (n=2) |
| No Symptoms | 3% (6/182) | 9% (n=18) | 4% (8/206) | 7% (n=14) |

#### 2.6 Reported Access to health care before death by period

Most of the deceased had access to hospital before death. Traditional medicine was almost absent. The access to hospital was similar in both periods. In the analysis by month we see that during the start of the pandemic, the access to hospital before death decrease temporarily.

Table 11. Reported health care seeking behaviours before death by period, Dagahaley refugee camp, Garissa County, Kenya

| | Pre-Pandemic | | Pandemic | |
|---|---|---|---|---|
| | Yes | Unknown | Yes | Unknown |
| Seek care | 83% (165/199) | 0% (n=1) | 75% (156/207) | 0% (n=0) |
| Hospital | 74% (147/199) | 0% (n=0) | 69% (142/207) | 0% (n=0) |
| Health center | 13% (26/196) | 2% (n=3) | 15% (31/206) | 0% (n=1) |
| Use Modern medicine | 2% (4/195) | 2% (n=4) | 0% (1/205) | 1% (n=2) |
| Self-Medication of Traditional medicine | 0% (0/196) | 2% (n=3) | 0% (0/206) | 0% (n=1) |
| Buying drug in the market | 1% (2/196) | 2% (n=3) | 0% (1/206) | 0% (n=1) |
| Traditional Practitionner | 0% (0/196) | 2% (n=3) | 0% (1/205) | 1% (n=2) |
| Pharmacist | 1% (2/195) | 2% (n=4) | 1% (2/205) | 1% (n=2) |

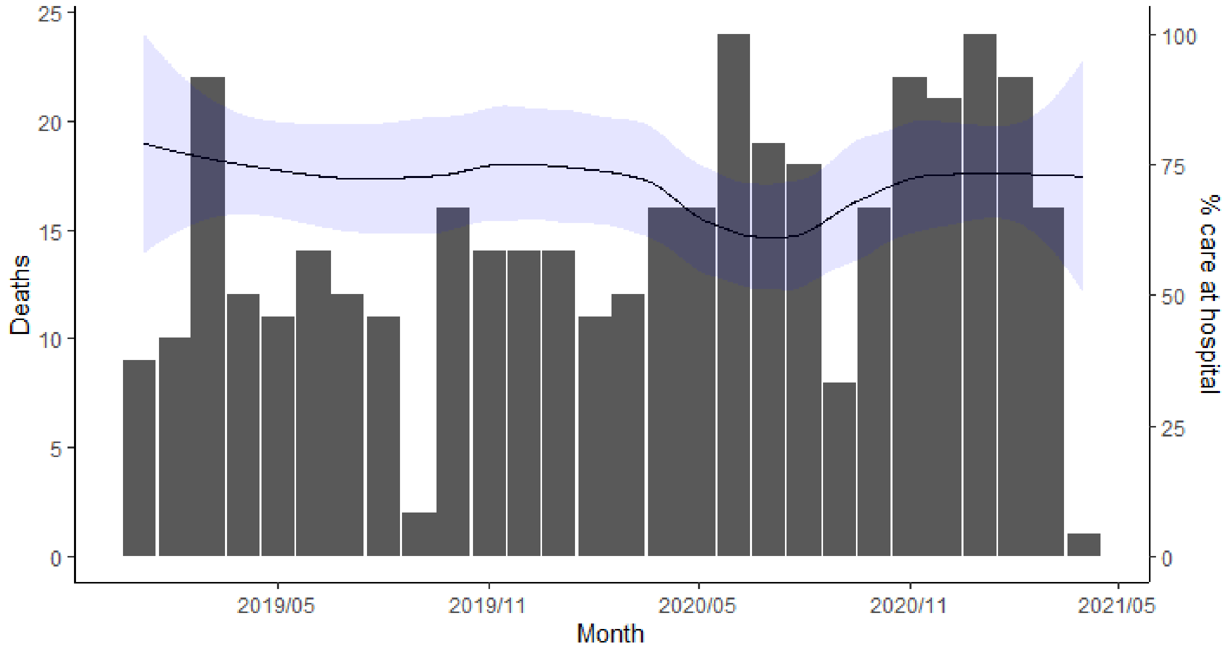

Figure 4. Number of deaths by month and Local polynomial Regression of the proportion of deceased that accessed the hospital before death, Dagahaley refugee camp, Garissa County, Kenya, May 2021

#### 2.7 Location of death by period

57% of the deceased died at home while 39% died in MSF hospital. theses proportions were similar in both periods.

Table 12. Reported place of death by period, Dagahaley refugee camp, Garissa County, Kenya

| | Pre-Pandemic | Pandemic | Total |
|---|---|---|---|
| Home | 54% (108) | 59% (122) | 57% (230) |
| On The way To Hospital | 2% (4) | 1% (3) | 2% (7) |
| MSF Hospital | 42% (84) | 36% (75) | 39% (159) |
| Other_Hospital | 2% (4) | 3% (7) | 3% (11) |
| Total | 100% (200) | 100% (207) | 100% (407) |

#### 2.8 Projection of COVID-19 related Death expected

Table 13. Expected Number of COVID-19 related deaths, Dagahaley refugee camp, Garissa County, Kenya

| Age group | Population | | | Seroprevalence | Nb infected | Infection Fatality Rate* | | Number of death |
|---|---|---|---|---|---|---|---|---|
| | Male | Female | Total | | | Male | Female | |
| 0-4 | 7923 | 7394 | 15317 | 1.09% | 166 | 0.003% | 0.003% | 0.005 |
| 5-9 | 7704 | 6957 | 14661 | 3.26% | 477 | 0.001% | 0.001% | 0.005 |
| 10-14 | 6611 | 5737 | 12348 | 5.43% | 670 | 0.001% | 0.001% | 0.007 |
| 15-19 | 4863 | 5610 | 10472 | 7.60% | 796 | 0.003% | 0.002% | 0.020 |
| 20-24 | 3588 | 3424 | 7012 | 8.68% | 609 | 0.008% | 0.005% | 0.040 |
| 25-29 | 2805 | 3715 | 6520 | 8.68% | 566 | 0.017% | 0.009% | 0.070 |
| 30-34 | 2459 | 2131 | 4590 | 8.68% | 399 | 0.033% | 0.015% | 0.098 |
| 35-39 | 1585 | 2186 | 3770 | 8.68% | 327 | 0.056% | 0.025% | 0.124 |
| 40-44 | 1730 | 1493 | 3224 | 8.68% | 280 | 0.106% | 0.044% | 0.216 |
| 45-49 | 1402 | 838 | 2240 | 8.68% | 195 | 0.168% | 0.073% | 0.258 |
| 50-54 | 1311 | 619 | 1931 | 8.68% | 168 | 0.291% | 0.123% | 0.397 |
| 55-59 | 346 | 364 | 710 | 8.68% | 62 | 0.448% | 0.197% | 0.197 |
| 60-64 | 583 | 565 | 1147 | 8.68% | 100 | 0.595% | 0.318% | 0.457 |
| 65-69 | 310 | 200 | 510 | 8.68% | 44 | 1.452% | 0.698% | 0.512 |
| 70-74 | 273 | 291 | 565 | 8.68% | 49 | 2.307% | 1.042% | 0.811 |
| 75-79 | 146 | 127 | 273 | 8.68% | 24 | 4.260% | 2.145% | 0.776 |
| 80-84 | 182 | 182 | 364 | 8.68% | 32 | 10.825% | 5.759% | 2.623 |
| 85-89 | 36 | 55 | 91 | 8.68% | 8 | 10.825% | 5.759% | 0.616 |
| 90-94 | 18 | 36 | 55 | 8.68% | 5 | 10.825% | 5.759% | 0.353 |
| 95-99 | 18 | 18 | 36 | 8.68% | 3 | 10.825% | 5.759% | 0.262 |
| **Total** | **43893** | **41944** | **85837** | **5,80 %** | **4978** | | | **7,847** |

* O'Driscol M, Ribeiro Dos Santos G, Wang L, Cummings DA, Azman AS, Paireau J, et al. Age-specific mortality and immunity patterns of SARS-CoV-2 infection in 45 countries. medRxiv [Internet]. 2020;2020.08.24.20180851. Available from: https://doi.org/10.1101/2020.08.24.20180851

##### 2.9 Expected mortality rate based on Kenyan mortality rates

Table 14. Expected mortality rate based on Kenyan mortality ratess , Dagahaley refugee camp, Garissa County, Kenya

| Age group | Population | | | Yearly risk of death* | | Number of Deaths /year | Crude Mortality rate per 10 000 pers per day |
|---|---|---|---|---|---|---|---|
| | Male | Female | Total | Male | Female | | |
| 0-4 | 7923 | 7394 | 15317 | 0.0114 | 0.0092 | 158 | 0.28 |
| 5-9 | 7704 | 6957 | 14661 | 0.002 | 0.001 | 22 | 0.04 |
| 10-14 | 6611 | 5737 | 12348 | 0.001 | 0.001 | 12 | 0.03 |
| 15-19 | 4863 | 5610 | 10472 | 0.002 | 0.001 | 15 | 0.04 |
| 20-24 | 3588 | 3424 | 7012 | 0.003 | 0.002 | 18 | 0.07 |
| 25-29 | 2805 | 3715 | 6520 | 0.004 | 0.003 | 22 | 0.09 |
| 30-34 | 2459 | 2131 | 4590 | 0.004 | 0.003 | 16 | 0.10 |
| 35-39 | 1585 | 2186 | 3770 | 0.006 | 0.004 | 18 | 0.13 |
| 40-44 | 1730 | 1493 | 3224 | 0.007 | 0.005 | 20 | 0.17 |
| 45-49 | 1402 | 838 | 2240 | 0.008 | 0.006 | 16 | 0.20 |
| 50-54 | 1311 | 619 | 1931 | 0.011 | 0.007 | 19 | 0.27 |
| 55-59 | 346 | 364 | 710 | 0.013 | 0.009 | 8 | 0.30 |
| 60-64 | 583 | 565 | 1147 | 0.02 | 0.014 | 20 | 0.47 |
| 65-69 | 310 | 200 | 510 | 0.03 | 0.023 | 14 | 0.75 |
| 70-74 | 273 | 291 | 565 | 0.047 | 0.038 | 24 | 1.16 |
| 75-79 | 146 | 127 | 273 | 0.075 | 0.064 | 19 | 1.91 |
| 80-84 | 182 | 182 | 364 | 0.122 | 0.108 | 42 | 3.15 |
| 85-89 | 36 | 55 | 91 | 0.226 | 0.205 | 19 | 5.84 |
| 90-94 | 18 | 36 | 55 | 0.226 | 0.205 | 12 | 5.80 |
| 95-99 | 18 | 18 | 36 | 0.226 | 0.205 | 8 | 5.90 |
| **Total** | **43893** | **41944** | **85837** | | | **502** | **0.16** |

* The Yearly risk of death by age group and sex in Kenya in 2016 (source WHO).

### 3 Data from Health facilities

##### 3.1 Consultation in MSF Primary health centres

From January 2019 to April 2020, 164 848 new consultations were reported in the two MSF health centre in the camp (N°4 and N°7), on average 339 consultations per day. From May 2020 to March 2021, 82187 new consultation were reported on average 245 per days, a decrease of 37% compared to pre pandemic period. The distribution of consultation per month (Figure 5) shows that the decrease started in April 2020, when awareness and mitigation measure were already set and  before the first case of COVID-19 was reported in Dagahaley (May 16th, 2020), and this decrease hasn’t come back to pre-pandemic level since.

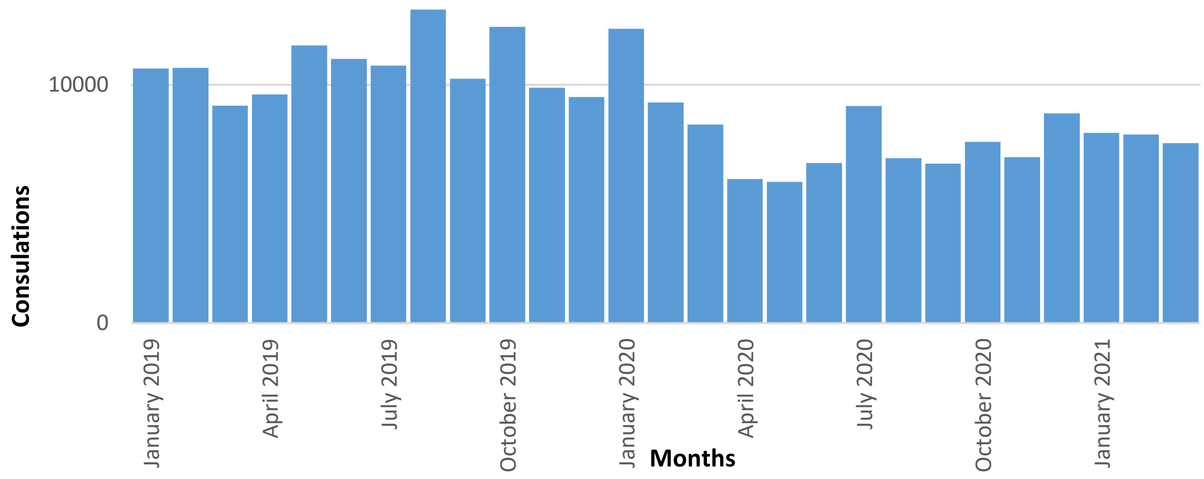

Figure 5. New Consultation in MSF Health Post N°4 and N°7 , Dagahaley refugee camp, Garissa County, Kenya, January 2019 to March 2021

#### 3.2 Consultation at the emergency room in MSF hospital

From April 2019 to April 2020, 46 105 consultations were reported in the Emergency room, on average 116 consultations per day. From May 2020 to March 2021, 35 961 new consultation were reported on average 107 per days, a slight decrease of 8% compared to pre pandemic period. the decrease is much less than in the health posts, it was strong at the start of the pandemic but came back to previous level in October 2020.

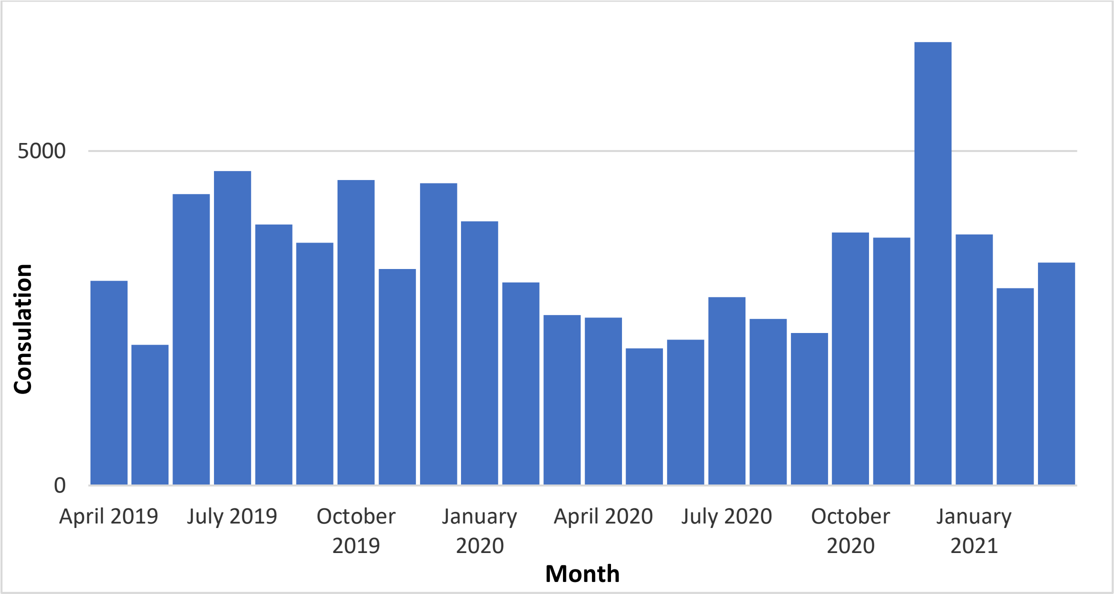

Figure 6. Consultation in Emergency room at MSF hospital, Dagahaley refugee camp, Garissa County, Kenya, April 2019 to March 2021

##### 3.3 Admission in MSF hospital

From January 2019 to April 2020, 12 918 admissions were reported in MSF hospital in Dagahaley, on average 26,5 admissions per day. From May 2020 to March 2021, 6467 admissions were reported on average 19,3 per days, a decrease of 37% compared to pre pandemic period. The distribution of admission per month (Figure 5) shows a similar pattern as the distribution of consultations, though a marked decrease started later, in June 2020.

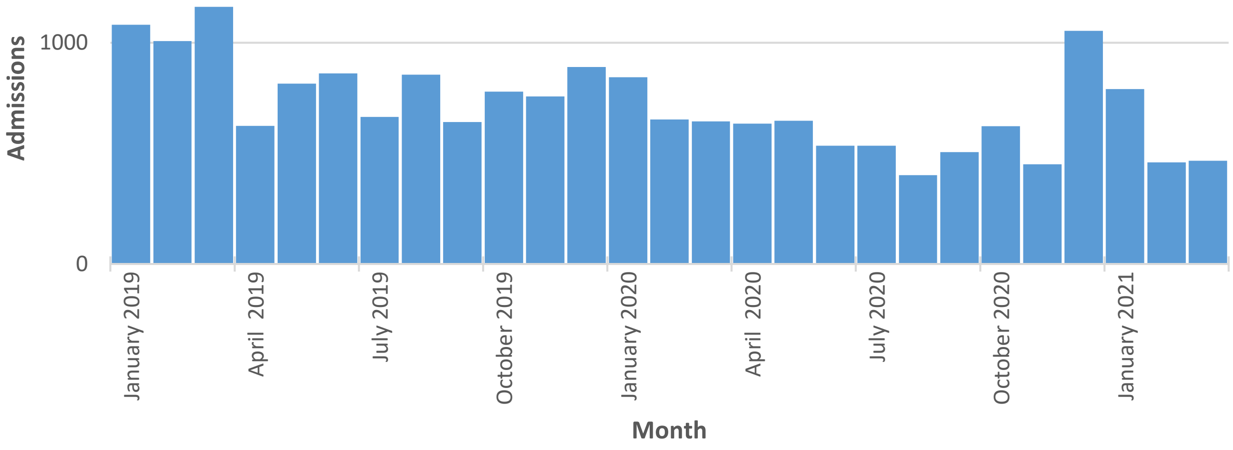

Figure 7. Admission in MSF Hospital, Dagahaley refugee camp, Garissa County, Kenya, January 2019 to March 2021

##### 3.4 Death in MSF hospital

From January 2019 to April 2020, 140 deaths from people living in the camp were registered in the Hospital mortality register, on average 0.288 death per day. From 1 May 2020 to 21 March 2021, 81 deaths were registered, on average 0.25 death per day, a 15 % decrease.

The distribution of death overtime doesn't show a specific pattern. The proportion of death of people over 50 since the start of the pandemic has slightly increased (figure 8).

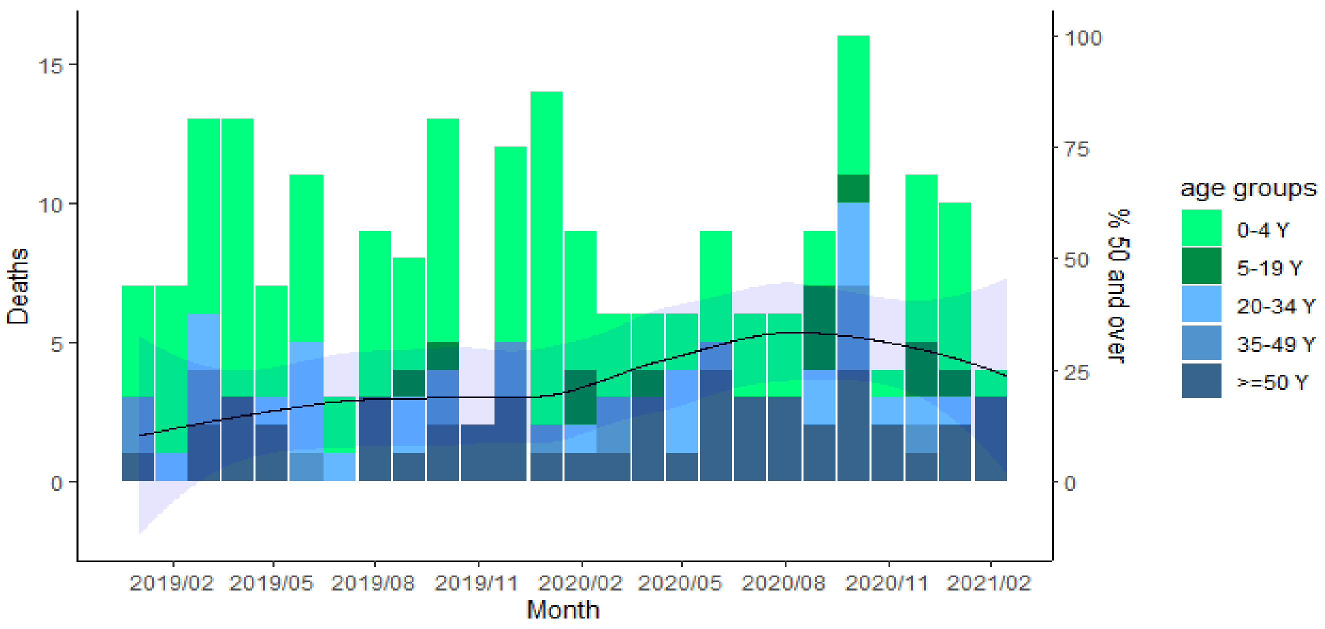

Figure 8. Death registered in MSF Hospital by month and local polynomial Regression of the proportion of death aged 50 years or more, Dagahaley refugee camp, Garissa County, Kenya, May 2021

### 4 Estimation of the Size of the Population in Dagahaley camp, MSF-OCG/Kenya/ Dagahaley Project (extract)

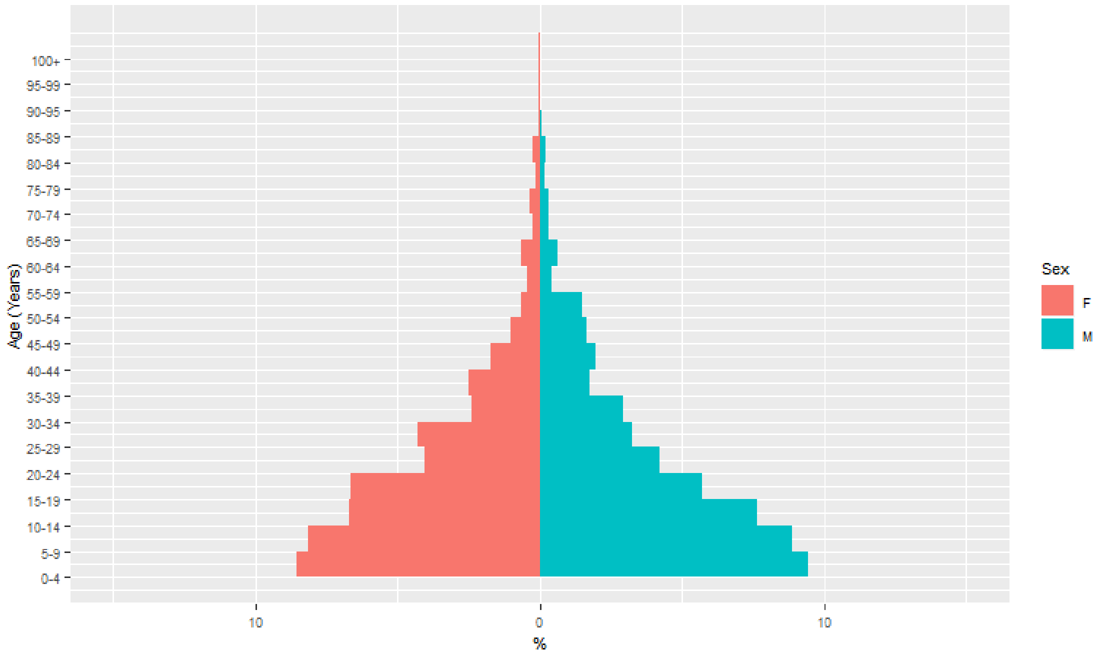

Figure 9. Age pyramid of the population of Dagahaley Camp, Kenya, 2018

Table 15. Age Distribution of the population of Dagahaley Kenya, 2018

| Age group | M | F | Total |
|---|---|---|---|
| 0-19 | 31.4% | 29.7% | 61.1% |
| 20-34 | 11.3% | 11.9% | 23.2% |
| 35-49 | 5.2% | 4.9% | 10.1% |
| >=50 | 3.1% | 2.5% | 5.7% |
